# Association of red cell distribution width to albumin ratio with risk of all-cause and cause-specific mortality: two prospective cohort studies

**DOI:** 10.1101/2024.01.10.24301123

**Authors:** Meng Hao, Shuai Jiang, Xiangnan Li, Shuming Wang, Yi Li, Jingdong Tang, Zixin Hu, Hui Zhang

**Affiliations:** Human Phenome Institute, Zhangjiang Fudan International Innovation Centre, Fudan University, Shanghai, 201203, China; Department of Vascular Surgery, Shanghai Key Laboratory of Vascular Lesion Regulation and Remodeling, Shanghai Pudong Hospital, Fudan University Pudong Medical Center, Shanghai, 201399, China; Artificial Intelligence Innovation and Incubation Institute, Fudan University, Shanghai, 200438, China

**Author notes:** Meng Hao and Shuai Jiang contributed equally to this work. Corresponding authors: Hui Zhang, Human Phenome Institute, Fudan University, Shanghai, 201203, China. E-mail address Zixi Hu, Artificial Intelligence Innovation and Incubation Institute, Fudan University, Shanghai, 200438, China. E-mail address Jingdong Tang, Department of Vascular Surgery, Shanghai Key Laboratory of Vascular Lesion Regulation and Remodeling, Shanghai Pudong Hospital, Fudan University Pudong Medical Center, Shanghai, 201399, China.

**Keywords:** red cell distribution width to albumin ratio, all-cause mortality, cause-specific mortality, cohort

## Abstract

**Background:** The red cell distribution width to albumin ratio (RAR) has emerged as a reliable prognostic marker for mortality in various diseases. However, whether RAR is associated with mortality remains unknown in the general population.

**Objective:** Explore whether RAR is associated with all-cause and cause-specific mortality, and to elucidate the dose-response relationship between them.

**Methods:** This study included 50622 participants aged 18+ years from US National Health and Nutrition Examination Survey (NHANES), and 418950 participants aged 37+ years from UK Biobank. The potential association between RAR and the risk of all-cause and cause-specific mortality was evaluated by Cox proportional hazard models. Restricted cubic spline regressions were applied to estimate the possible nonlinear relationships.

**Results:** NHANES documented 7590 deaths over a median follow-up of 9.4 years, and UK Biobank documented 36793 deaths over a median follow-up of 14.5 years. In multivariable analysis, elevated RAR were significantly associated with a higher risk of all-cause mortality (NHANES: hazard ratio [HR]: 1.86, 95% confidence interval [CI]: 1.81-1.93; UK Biobank: HR: 2.01, 95% CI: 1.96-2.06), as well as mortality due to malignant neoplasms, heart disease, cerebrovascular diseases, respiratory diseases, diabetes mellitus, and others in both the two cohorts (all P-value < 0.05).

**Conclusions:** Higher baseline RAR was strongly and independently associated with increased risk of all-cause and cause-specific mortality in the general population. RAR was a promising indicator that simply, reliably, and inexpensively accessible for identifying high-risk of mortality in clinical practice.

## Introduction

The identification of new biomarkers for early identification of increased risk of mortality remains an unmet need. There has been significant progress in establishing the epidemiology of novel blood biomarkers on all-cause and cause-specific mortality, such as serum neurofilament light chain[1, 2], N-terminal pro-brain natriuretic peptide[3], retinol[4], alpha-tocopherol[5], and beta carotene[6]. Despite these progresses, simple, inexpensive, and reliable indicators are still essential for predicting mortality in clinical practice. At present, the routine laboratory test is widely used in primary care and hospitals, and further utilization of routine clinical biomarkers is expected. For example, several new biomarkers were derived from routine laboratory test, such as prognostic nutritional index, neutrophil/lymphocyte ratio, and platelet/lymphocyte ratio. And these simply integrated biomarkers showed significant associations with all-cause and cause-specific mortality in the general population[7–9]. Therefore, there is still substantial interest in the use of routine biomarkers to identify persons who are at risk for mortality.

The red blood cell distribution width (RDW), a marker from routine laboratory test, reflects the degree of heterogeneity of erythrocyte volume. The increased levels of RDW, resulting from impaired erythropoiesis and abnormal red blood cell survival, is associated with a variety of disorders and mortality[10]. Serum albumin, the most abundant circulating protein in the blood, is an important marker of nutritional status and inflammatory response[11, 12]. Physiological properties of albumin include anti-inflammatory, antioxidant, anticoagulant and antiplatelet aggregation activity as well as colloid osmotic effect[13]. Several studies have reported that serum albumin was inversely associated with incidence of dysfunction, diseases, and mortality[14–16]. Additionally, RDW is positively correlated with chronological age, while serum albumin is negatively correlated with chronological age[17]. RDW and albumin were enrolled in the construction of phenotypic and biological age, and contributed a lot to individual variances of aging pace[18–20].

Both RDW and albumin are suggested as an integrative biomarker for a multidimensional dysfunctional physiological status that relates to inflammation, oxidative stress, and nutrition[10–12], while they depict these pathological aspects from different scopes. Therefore, considering their essential roles in physiological function, diseases, and assessment of aging, the integration of the two markers may be substantial valuable to predict mortality. The red cell distribution width to albumin ratio (RAR), a derived marker from routine laboratory test, may convey massive information beyond RDW and albumin. Most recently, RAR has emerged as a potential risk biomarker for adverse outcomes in various diseases, including acute myocardial infarction[21], atrial fibrillation[22], diabetes[23], heart failure[24].[25], chronic kidney disease[26], and stroke[27]. However, whether RAR is associated with mortality remains unknown in the general population.

Hence, to address this knowledge gap, we first hypothesized that elevated RAR is associated with an increased risk of mortality in the general population, and then examined the potential association between RAR levels and the risk of all-cause and cause-specific mortality using data from the US National Health and Nutrition Examination Survey (NHANES) and UK Biobank.

## METHODS

### Study population

The US NHANES is a nationally representative cross-sectional survey of civilian, noninstitutionalized persons living in the US. We used data from the NHANES 1988-2018. The data included demographic information, physical examination results, questionnaire items, and mortality status linked from the National Death Index. In this cohort, we used data from a publicly available data dictionary[28], and participants aged 18 years or older with complete data on serum albumin, RDW and death register were included and analyzed. The final sample included 50622 (aged 19-85 years) individuals. Because the data in our study were publicly available and performed as a secondary analysis, no additional ethical review was necessary.

The UK Biobank is a large-scale, population-based, and prospective cohort study consisting of over 500000 adults (aged 37-73 years) recruited across England, Scotland, and Wales. During the baseline period (2006 to 2010), information about participants were collected through questionnaires, physical measurements, and biological samples. All participants provided electronic informed consent. The UKB study received approval from the National Information Governance Board for Health and Social Care and the National Health Service North West Multi-Centre Research Ethics Committee. In this cohort, a total of 418950 participants with complete data on serum albumin, RDW and death register were included and analyzed.

### Measurement of the RAR

In NHANES, serum albumin was determined using a bromocresol purple method. The RDW (%) was measured by the Coulter analyzer in the Mobile Examination Centers using peripheral blood. In UK Biobank, serum albumin was measured using a colorimetric approach and a Beckman Coulter AU5800 assay (Beckman Coulter, United Kingdom). The RDW (%) was measured using four Beckman Coulter LH750 instruments within 24 hours of blood draw, with extensive quality control performed by UK Biobank. In our study, the independent variable was RAR, which was defined as the ratio of RDW/albumin.

### Outcomes

In this study, the primary outcome was all-cause mortality. The secondary outcomes cause-specific mortality. In US NHANES, dates and causes of death were linked to the National Death Index records to 31 December 2019. In UK Biobank, dates and causes of death were obtained from the National Health Service Information Centre (England and Wales) and the National Health Service Central Register Scotland (Scotland) to 30 November 2022. We used the 10th revision of the International Statistical Classification of Diseases (ICD-10) to identify causes of death. Mortality outcomes in this study included the following underlying causes of death: malignant neoplasms, heart diseases, cerebrovascular diseases, respiratory diseases, Alzheimer’s disease, and others.

### Covariates

In this study, covariates included demographic characteristics, lifestyle factors, and clinical information. Demographic characteristics included age at baseline, gender, race/ethnicity, and education. Lifestyle factors included body mass index (BMI), smoking status and alcohol drinking. BMI was used to divide participants into underweight (< 18 kg/m^2^) normal (18-25 kg/m^2^), overweight (25-30 kg/m^2^), and obese groups (> 30 kg/m^2^). Clinical information was obtained from self-reports by participants or proxies, including the presence of hypertension, diabetes mellitus, heart disease, stroke, and cancer.

### Statistical Analysis

Frist, we describe data from the two samples with the mean and standard deviation (SD) or frequency (%) for continuous and categorical variables, respectively.

Second, we utilized Cox proportional hazard models to estimate the hazard ratios (HR) and 95% confidence intervals (CI) of continuous RAR levels associated with all-cause and cause-specific mortality after adjusting for age, gender, education, race, smoking status, BMI category, hypertension, diabetes, heart disease, stroke, and cancer.

Third, subgroups were created according to age groups (< 45, 45-64, and 65+ years), gender (male and female), BMI category (underweight, normal, overweight, and obese), race/ethnicity (White and non-White), smoking status (yes and no), and alcohol drinking (yes and no). Stratified analysis was also conducted to examine the association between levels of RAR and all-cause mortality.

Forth, we conducted restricted cubic spline regressions with 3-knots located at the 10th, 50th, and 90th percentiles of RAR to estimate possible non-linear between RAR and the risk of all-cause and cause-specific mortality with adjusted for confounder factors.

Last, we categorized the RAR level into four groups according to quartile (for NHANES, Q1: 2.02-2.82, Q2: 2.82-3.05, Q3: 3.05-3.35, Q4: 3.35-12.1; for UK Biobank, Q1: 0.46-2.80, Q2: 2.80-2.96, Q3: 2.96-3.14, Q4: 3.14-10.1), and then examined the association between the RAR groups and risk of mortality. We also conducted Kaplan‒Meier survival to plot the association of different RAR groups with all-cause and cause-specific mortality.

All results were considered significant at a P-value < 0.05 (2-tailed). All analyses were conducted using R statistical software (version 4.2.1; www.r-project.org).

## RESULTS

### Characteristics of the study population

**Table 1** shows the characteristics of the study population in NHANES and UK Biobank. Baseline demographic characteristics, lifestyle factors, and chronic diseases of the study population are presented in detail. At baseline, a total of 50622 (48.37% males, 51.63% females) participants in NHANES and 418950 (46.29% males, 53.71% females) participants in UK Biobank were analyzed. The mean age of participants were 48.57 (18.72) years in NHANES and 56.56 (8.09) in UK Biobank, respectively. The mean levels of RAR were 3.15 (0.51) in NHANES and 2.99 (0.31) in UK Biobank, respectively. In addition, NHANES documented 7590 deaths over a median follow-up of 9.4 years, and UK Biobank documented 36793 deaths over a median follow-up of 14.5 years.

**Table 1.** Baseline characteristics of study population.

|  | NHANES (N = 50622) | UK Biobank (N = 418950) |
| --- | --- | --- |
| Age, years, M (SD) | 48.57 (18.72) | 56.56 (8.09) |
| Age category, N (%) |  |  |
| < 45, years | 22716 (44.87%) | 42897 (10.24%) |
| 45-64, years | 15878 (31.37%) | 295704 (70.58%) |
| 65+, years | 12028 (23.76%) | 80349 (19.18%) |
| Gender, N (%) |  |  |
| Men | 24486 (48.37%) | 193912 (46.29%) |
| Women | 26136 (51.63%) | 225038 (53.71%) |
| Race, N (%) |  |  |
| White | 22277 (44.01%) | 370147 (88.35%) |
| Non-White | 23749 (55.99%) | 48803 (11.65%) |
| Educational status, N (%) |  |  |
| College or above | 24331 (48.06%) | 135232 (32.31%) |
| High school or equivalent | 11270 (22.26%) | 207086 (49.48%) |
| Less than high school | 13181 (26.04%) | 71628 (17.11%) |
| Alcohol drinking, N (%) |  |  |
| Yes | 32792 (66.64%) | 384483 (91.87%) |
| No | 12701 (25.81%) | 33422 (7.99%) |
| Smoking status, N (%) |  |  |
| Ever | 22369 (45.45%) | 44129 (10.54%) |
| Never | 26802 (54.46%) | 372703 (89.05%) |
| BMI, kg/m<sup>2</sup>, M (SD) | 28.86 (6.78) | 27.43 (4.78) |
| BMI categories, N (%) |  |  |
| Underweight (<18.5) | 847 (1.70%) | 2141 (0.51%) |
| Normal (18.5-25) | 14311 (28.77%) | 135349 (32.44%) |
| Overweight (25-30) | 16701 (33.57%) | 177934 (42.64%) |
| Obese (≥ 30) | 17888 (35.96%) | 101857 (24.41%) |
| RDW, %, M (SD) | 13.17 (1.34) | 13.49 (0.98) |
| Albumin, g/dL, M (SD) | 4.23 (0.37) | 4.52 (0.26) |
| RAR, %/(g/dL), M (SD) | 3.15 (0.51) | 2.99 (0.31) |
| Chronic diseases, N (%) |  |  |
| Hypertension | 2054 (4.05%) | 100408 (23.99%) |
| Diabetes | 5896 (11.65%) | 21821 (5.21%) |
| Heart disease | 3808 (7.80%) | 9664 (2.31%) |
| Stroke | 1835 (3.76%) | 5114 (1.22%) |
| Cancer | 4423 (9.05%) | 31583 (7.55%) |

RAR: red cell distribution width to albumin ratio, RDW: red cell distribution width, BMI: body mass index.

### Association between RAR and mortality

The results of Cox proportional hazard analyses regarding the association of RAR with the risk of all-cause and cause-specific mortality are presented in **Table 2**. In NHANES, after adjusting for covariates, elevated RAR levels were associated with increased risk of all-cause mortality (hazard ratio [HR]: 1.86, 95% confidence interval [CI]: 1.81-1.93), malignant neoplasms (HR: 1.98, 95% CI: 1.82-2.16), heart diseases (HR: 1.99, 95% CI: 1.84-2.15), cerebrovascular diseases (HR: 1.51, 95% CI: 1.21-1.88), respiratory diseases (HR: 1.93, 95% CI: 1.64-2.27), Alzheimer’s disease (HR: 1.36, 95% CI: 1.83), diabetes mellitus (HR: 1.63, 95% CI: 1.34-1.99), others caused (HR: 2.00, 95% CI: 1.90-2.12) mortality. In UK Biobank, similar positive associations were also found between RAR levels and all-cause mortality (HR: 2.01, 95% CI: 1.96-2.06), malignant neoplasms (HR: 1.86, 95% CI: 1.79-1.93), heart diseases (HR: 2.32, 95% CI: 2.19-2.46), cerebrovascular diseases (HR: 2.06, 95% CI: 1.83-2.32), respiratory diseases (HR: 2.80, 95% CI: 2.63-2.99), diabetes mellitus (HR: 2.73, 95% CI: 2.28-3.27), others caused (HR: 2.33, 95% CI: 2.24-2.43) mortality.

**Table 2.** Hazard ratios of all-cause and cause-specific mortality according to continuous levels of RAR.

|  |  | Model 1 |  |  | Model 2 |  |
| --- | --- | --- | --- | --- | --- | --- |
|  | **Event** | **HR (95% CI)** | **P-value** | **Event** | **HR (95% CI)** | **P-value** |
| **NHANES** |  |  |  |  |  |  |
| All-cause Mortality | 7590 | 2.01 (1.96-2.07) | < 0.001 | 6605 | 1.86 (1.8-1.93) | < 0.001 |
| Underlying cause |  |  |  |  |  |  |
| Malignant neoplasms | 1638 | 1.97 (1.84-2.10) | < 0.001 | 1476 | 1.98 (1.82-2.16) | < 0.001 |
| Heart diseases | 1961 | 2.17 (2.05-2.29) | < 0.001 | 1717 | 1.99 (1.84-2.15) | < 0.001 |
| Cerebrovascular diseases | 433 | 1.88 (1.64-2.15) | < 0.001 | 357 | 1.51 (1.21-1.88) | < 0.001 |
| Respiratory diseases | 412 | 2.07 (1.82-2.35) | < 0.001 | 367 | 1.93 (1.64-2.27) | < 0.001 |
| Alzheimer's disease | 283 | 1.70 (1.41-2.06) | < 0.001 | 240 | 1.36 (1.00-1.83) | 0.048 |
| Diabetes mellitus | 277 | 2.21 (1.91-2.55) | < 0.001 | 246 | 1.63 (1.34-1.99) | < 0.001 |
| Others | 2586 | 2.13 (2.05-2.23) | < 0.001 | 2202 | 2.00 (1.90-2.12) | < 0.001 |
| **UK Biobank** |  |  |  |  |  |  |
| All-cause Mortality | 36793 | 2.32 (2.28-2.37) | < 0.001 | 35843 | 2.01 (1.96-2.06) | < 0.001 |
| Underlying cause |  |  |  |  |  |  |
| Malignant neoplasms | 17657 | 2.14 (2.08-2.21) | < 0.001 | 17323 | 1.86 (1.79-1.93) | < 0.001 |
| Heart diseases | 5131 | 2.57 (2.46-2.68) | < 0.001 | 4976 | 2.32 (2.19-2.46) | < 0.001 |
| Cerebrovascular diseases | 1623 | 2.36 (2.17-2.58) | < 0.001 | 1574 | 2.06 (1.83-2.32) | < 0.001 |
| Respiratory diseases | 2636 | 2.85 (2.71-2.99) | < 0.001 | 2555 | 2.80 (2.63-2.99) | < 0.001 |
| Alzheimer's disease | 687 | 1.34 (1.06-1.68) | 0.013 | 675 | 0.87 (0.66-1.15) | 0.331 |
| Diabetes mellitus | 253 | 3.19 (2.83-3.60) | 0.002 | 243 | 2.73 (2.28-3.27) | < 0.001 |
| Other mortality | 8806 | 2.51 (2.43-2.60) | < 0.001 | 8479 | 2.33 (2.24-2.43) | < 0.001 |

Model 1: Crude model;
Model 2: Adjusted for age, gender, body mass index, race, educational levels, smoking status, alcohol drinking, hypertension, diabetes, heart disease, stroke, and cancer.

In addition, we conducted stratified analyses in the two cohorts across various subgroups according to age groups, gender, BMI category, race/ethnicity, smoking status, and alcohol drinking (**Figure 1**). We found significant associations of elevated RAR levels with increased risk of all-cause mortality in each stratum (P-value < 0.05). These findings suggested that elevated RAR levels with increased risk of all-cause mortality, regardless of socioeconomic status, lifestyle factors, chronic diseases, and mental health. We also used a restricted 3-knot cubic spline regression to evaluate the RAR-mortality dose-response relation, with modeling RAR as a continuous variable (**Figure 2**). We found linear positive relationship between RAR with all-cause and cause-specific mortality in both the two cohorts (P for nonlinear < 0.05).

**Figure 1.**
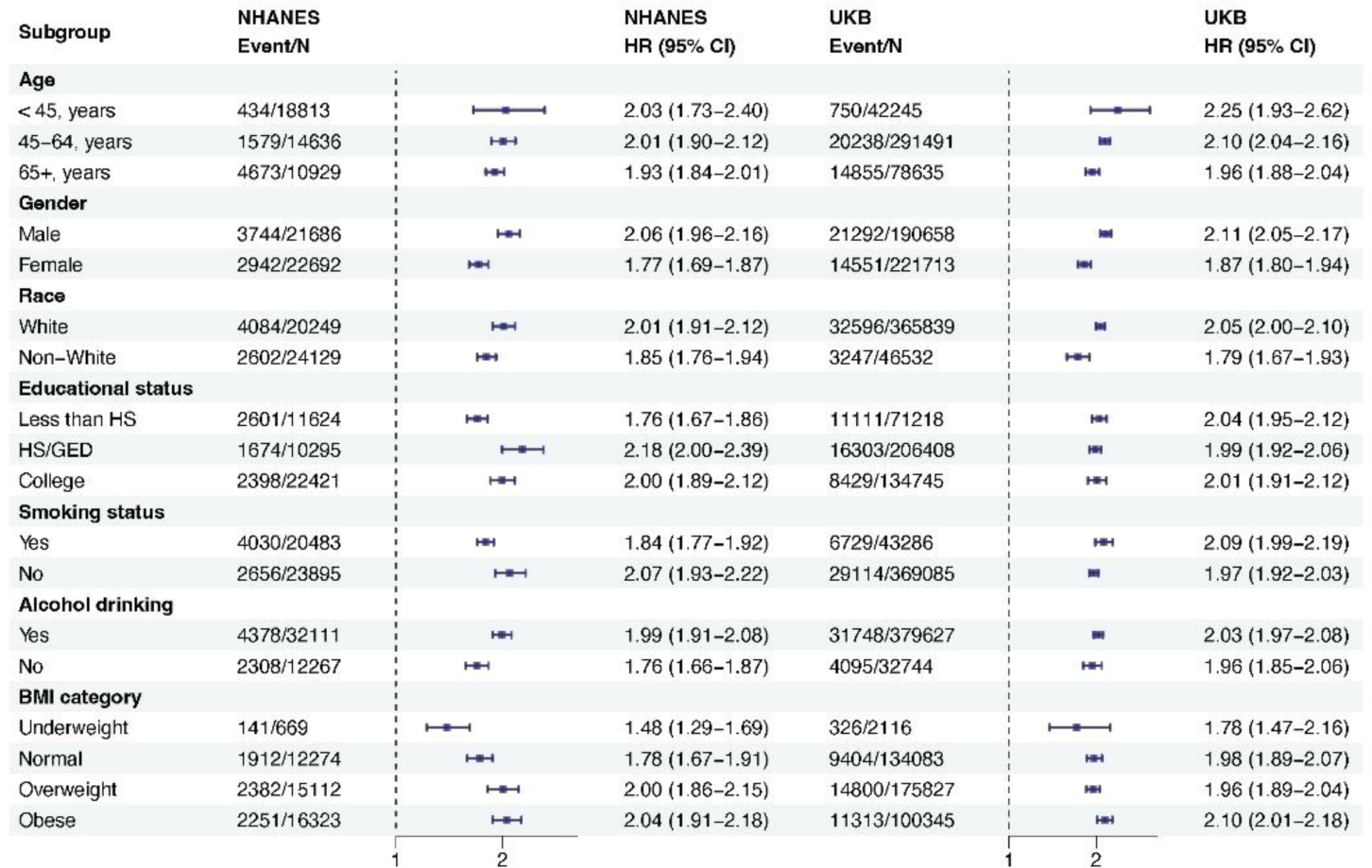
Association between RAR and all-cause mortality among subgroups. All models were adjusted for age, gender, body mass index, race, educational levels, smoking status, drinking status, hypertension, diabetes, heart disease, stroke, and cancer.

**Figure 2.**
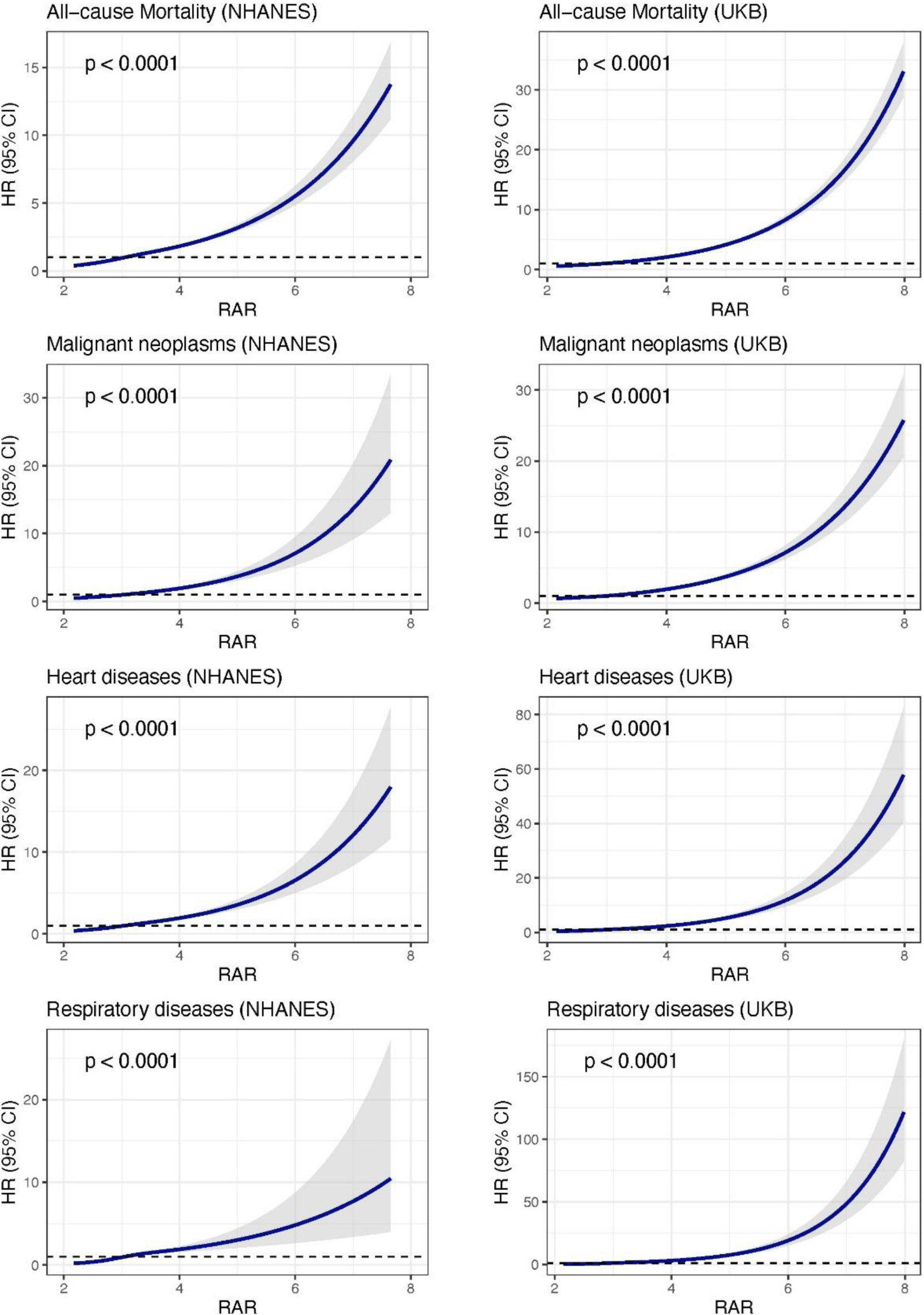
Cubic spline regression for estimated hazard ratios of mortality according to continuous levels of RAR. All models were adjusted for age, gender, body mass index, race, educational levels, smoking status, drinking status, hypertension, diabetes, heart disease, stroke, and cancer.

In further analyses, we categorized the RAR level into four groups according to quartile in NHANES and UK Biobank, and then examined the association between the RAR groups and risk of mortality (**Table 3**). Compared with participants in the lowest quintile of RAR, those in the higher quintiles experienced significantly higher mortality from all-cause, malignant neoplasms, heart diseases, cerebrovascular diseases, respiratory diseases, diabetes mellitus, others caused mortality, representing 7% to 340% risk increment (all P-trend < 0.0001). Kaplan-Meier survival plots demonstrate that participants in the higher quintile of RAR had significantly increased cumulative all-cause mortality than those in the lowest quintiles (P-value<0.0001, **Figure 3**), and similar patterns were observed for cause-specific mortality (P-value<0.0001, **Figure 3**).

**Figure 3.**
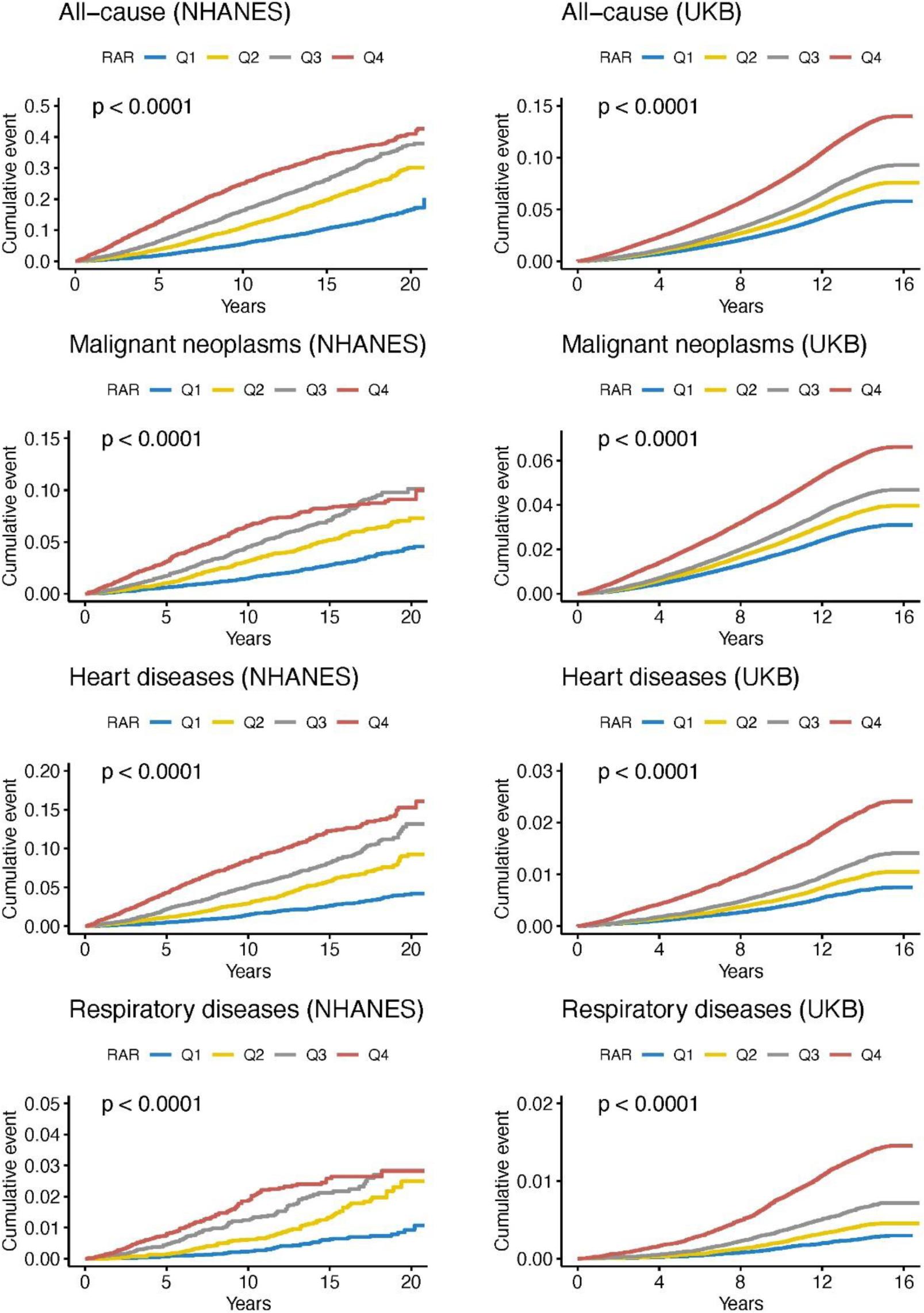
Kaplan-Meier curves of all-cause and cause-specific mortality according to quintiles of RAR.

**Table 3.** Hazard ratios of all-cause and cause-specific mortality according to quartile of RAR.

|  | NHANES |  |  | UK Biobank |  |  |
| --- | --- | --- | --- | --- | --- | --- |
|  | Event | HR (95% CI) | P-value | Event | HR (95% CI) | P-value |
| All-cause |  |  |  |  |  |  |
| Q1 | 1038 | 1.00 (ref) | 1.00 (ref) | 5712 | 1.00 (ref) | 1.00 (ref) |
| Q2 | 1578 | 1.17 (1.08-1.27) | < 0.001 | 7481 | 1.12 (1.08-1.16) | < 0.001 |
| Q3 | 1831 | 1.45 (1.33-1.57) | < 0.001 | 9051 | 1.22 (1.18-1.26) | < 0.001 |
| Q4 | 2158 | 2.26 (2.08-2.45) | < 0.001 | 13599 | 1.73 (1.68-1.79) | < 0.001 |
| P for trend |  |  | < 0.001 |  |  | < 0.001 |
| Malignant neoplasms |  |  |  |  |  |  |
| Q1 | 264 | 1.00 (ref) | 1.00 (ref) | 3017 | 1.00 (ref) | 1.00 (ref) |
| Q2 | 355 | 1.07 (0.91-1.26) | 0.43 | 3797 | 1.09 (1.04-1.14) | < 0.001 |
| Q3 | 415 | 1.39 (1.18-1.64) | < 0.001 | 4436 | 1.17 (1.12-1.23) | < 0.001 |
| Q4 | 442 | 2.03 (1.70-2.42) | < 0.001 | 6073 | 1.56 (1.49-1.63) | < 0.001 |
| P for trend |  |  | < 0.001 |  |  | < 0.001 |
| Heart diseases |  |  |  |  |  |  |
| Q1 | 245 | 1.00 (ref) | 1.00 (ref) | 692 | 1.00 (ref) | 1.00 (ref) |
| Q2 | 394 | 1.13 (0.96-1.33) | 0.13 | 964 | 1.18 (1.07-1.30) | < 0.001 |
| Q3 | 473 | 1.47 (1.25-1.74) | < 0.001 | 1259 | 1.39 (1.26-1.52) | < 0.001 |
| Q4 | 605 | 2.37 (2.01-2.80) | < 0.001 | 2061 | 2.12 (1.94-2.32) | < 0.001 |
| P for trend |  |  | < 0.001 |  |  | < 0.001 |
| Cerebrovascular diseases |  |  |  |  |  |  |
| Q1 | 61 | 1.00 (ref) | 1.00 (ref) | 245 | 1.00 (ref) | 1.00 (ref) |
| Q2 | 108 | 1.11 (0.81-1.53) | 0.52 | 342 | 1.17 (0.99-1.38) | 0.06 |
| Q3 | 95 | 1.11 (0.78-1.56) | 0.56 | 411 | 1.26 (1.07-1.48) | 0.0047 |
| Q4 | 93 | 1.57 (1.08-2.29) | 0.02 | 576 | 1.70 (1.46-1.99) | < 0.001 |
| P for trend |  |  | 0.049 |  |  | < 0.001 |
| Respiratory diseases |  |  |  |  |  |  |
| Q1 | 46 | 1.00 (ref) | 1.00 (ref) | 265 | 1.00 (ref) | 1.00 (ref) |
| Q2 | 92 | 1.49 (1.04-2.15) | 0.03 | 418 | 1.32 (1.13-1.54) | < 0.001 |
| Q3 | 118 | 2.05 (1.43-2.96) | < 0.001 | 644 | 1.75 (1.51-2.02) | < 0.001 |
| Q4 | 111 | 3.54 (2.39-5.25) | < 0.001 | 1228 | 3.22 (2.81-3.69) | < 0.001 |
| P for trend |  |  | < 0.001 |  |  | < 0.001 |
| Alzheimer's disease |  |  |  |  |  |  |
| Q1 | 48 | 1.00 (ref) | 1.00 (ref) | 148 | 1.00 (ref) | 1.00 (ref) |
| Q2 | 65 | 0.87 (0.59-1.28) | 0.48 | 158 | 0.84 (0.67-1.05) | 0.13 |
| Q3 | 78 | 1.11 (0.76-1.63) | 0.60 | 177 | 0.84 (0.67-1.05) | 0.12 |
| Q4 | 49 | 1.28 (0.82-1.98) | 0.28 | 192 | 0.88 (0.71-1.10) | 0.27 |
| P for trend |  |  | 0.192 |  |  | 0.314 |
| Diabetes mellitus |  |  |  |  |  |  |
| Q1 | 37 | 1.00 (ref) | 1.00 (ref) | 21 | 1.00 (ref) | 1.00 (ref) |
| Q2 | 35 | 0.74 (0.46-1.19) | 0.22 | 38 | 1.69 (0.99-2.88) | 0.05 |
| Q3 | 74 | 1.56 (1.01-2.40) | 0.04 | 52 | 2.14 (1.28-3.61) | 0.004 |
| Q4 | 100 | 2.32 (1.51-3.58) | < 0.001 | 132 | 4.40 (2.73-7.10) | < 0.001 |
| P for trend |  |  | < 0.001 |  |  | < 0.001 |
| Others |  |  |  |  |  |  |
| Q1 | 337 | 1.00 (ref) | 1.00 (ref) | 1324 | 1.00 (ref) | 1.00 (ref) |
| Q2 | 529 | 1.28 (1.11-1.47) | < 0.001 | 1764 | 1.15 (1.07-1.24) | < 0.001 |
| Q3 | 578 | 1.50 (1.30-1.74) | < 0.001 | 2072 | 1.24 (1.15-1.33) | < 0.001 |
| Q4 | 758 | 2.71 (2.34-3.13) | < 0.001 | 3337 | 1.94 (1.81-2.07) | < 0.001 |
| P for trend |  |  | < 0.001 |  |  | < 0.001 |

All models were adjusted for age, gender, body mass index, race, educational levels, smoking status, alcohol drinking, hypertension, diabetes, heart disease, stroke, and cancer.

## DISCUSSION

To our knowledge, this is the first and largest study to examine the association of RAR with all-cause and cause-specific mortality. Using data from the NHANES and UK Biobank, which involving approximately 470000 participants at baseline and 15000 deaths during follow-up periods, we found that higher levels of RAR were strongly and independently associated with increased risk of all-cause and cause-specific (including malignant neoplasms, heart disease, cerebrovascular diseases, respiratory diseases, diabetes mellitus, and others) mortality in the general population. Our findings from the restricted cubic spline analysis also supported increasing all-cause and cause-specific mortality with increasing RAR levels. In addition, both RDW and serum albumin were simply, reliably, and inexpensively accessible biomarkers from routine laboratory test. Hence, the RAR could be recognized as a promising indicator for identifying high-risk of mortality in clinical practice.

Although our study is the first to examine the association of RAR with mortality in the general population, previous studies have investigated their relationships in various diseases-specific populations, such as patients with acute myocardial infarction[21], atrial fibrillation[22], diabetes mellitus[23], heart failure[24]. [25], stroke[27], burn surgery[29], cervical cancer[30]. For example, Hong et.al included 860 patients with diabetic foot ulcers in China, and found a positive association between high RDR and all-cause mortality (HR 2.36, 95% CI: 1.41-3.94) after adjusted cofounder factors[23]. Zhang et.al included 22672 and 60754 patients with chronic heart failure from the Medical Information Mart for Intensive Care Database iii V1.4 (MIMIC-III) and NHANES, respectively. They found that elevated levels of RAR were significant associated with all-cause mortality both MIMIC (HR 1.12, 95% CI: 1.1-1.14) and NHANES (HR 2.26, 95% CI: 1.52-3.36)[31]. Zhao et.al included 1480 patients with stroke from the MIMIC-III, and reported that patients in the highest quartile of RAR had the highest 30-day (HR1.88, 95% CI: 1.36-2.58), 90-day (HR 2.12, 95% CI: 1.59-2.82), and one-year (HR 2.15, 95% CI: 1.65-2.80) all-cause mortality, compared to those in the first quartile[27].

In addition, RAR has also emerged as a reliable prognostic marker for other adverse outcomes in various diseases at present. Kimura et.al conducted a retrospective cohort study using 997 chronic kidney disease patients who were enrolled in the Fukushima Cohort Study[26]. They found that patients in the highest tertile of RAR showed a significantly higher risk of end-stage kidney disease (HR 2.92, 95% CI: 1.44-5.94), all-cause mortality (HR 3.38, 95% CI: 1.81-6.30) and cardiovascular events (HR 2.27, 95% CI: 1.36-3.78), than those in the lowest tertile of RAR. Huang et.al included and analyzed 10267 patients with coronary heart disease in China[32]. After adjusting for confounding factors, they found that elevated RAR was significantly associated with increased risk of carotid plaque formation (OR: 1.23; 95% CI 1.08-1.39), and the association was more significant in women and patients aged < 60 years[32]. In summary, previous studies have reported that RAR was a potential risk biomarker for adverse outcomes in various diseases-specific populations. While we examined the association of RAR levels with all-cause and cause-specific mortality in the general population, and revealed positive and independent associations using data from two large population-based cohorts with longer follow-up periods. Our finding provided strong epidemiologic support to the association that elevated RAR levels were associated with increased risk of all-cause and cause-specific mortality in the general population.

### Biological mechanisms

The biological mechanisms underlying the association of higher RAR with mortality risk is unclear. It seems likely that these findings are due to chronic inflammation and nutritional status. As we known, chronic inflammation and nutritional status played important role in development of mortality[7, 33], and various chronic diseases, including cardiovascular disease, cancer, diabetes mellitus, chronic kidney disease, and neurodegenerative disorders[34]. In detail, RDW reflects the degree of heterogeneity of erythrocyte volume. The increased levels of RDW could mirror impaired erythropoiesis and abnormal red blood cell survival through a variety of underlying metabolic abnormalities, such as inflammation and poor nutritional status[10]. Serum albumin is an important marker of nutritional status and inflammatory response[11, 12]. Previous studies have reported that higher RDW[35–38] and lower albumin[14–16] were both associated with increased risk of incident chronic diseases and mortality in the general population. Therefore, we could hypothesis that the combined of the two markers could simultaneously reflect the inflammation and nutritional status in lifespan, and then related to all-cause and cause-specific mortality.

### Strengths and limitations

Our study has several strengths. We conducted this study in two large, prospective, and population-based cohorts with a long follow-up period for cause-specific mortality. The current study has ability to obtain relatively robust results with high statistical efficiency. Additionally, we used data from UK Biobank and NHANES, which enrolled the general population with various racial background and geographic location. The findings in our study may be more generalizable. Several limitations of the study also deserve mention. First, this study was an observational study, the association of RAR with all-cause and cause-specific mortality is merely correlational rather than a causal relationship. Second, the levels of RAR were assessed only at a one-time point, and its potential trajectories were not accounted for in this study. The single measurement of RAR might lead to some misclassification in these two populations. Further studies with repeated measurements of RAR should be conducted to capture its trajectories, and then validated our findings in the future. Last, although we have controlled for many potential confounders, including demographics factors, chronic disease, and lifestyles, we cannot rule out all the potential residual confounding, especially by unmeasured variables.

## CONCLUSION

In conclusion, the RAR, which deprived from blood RDW and serum albumin, was strongly and independently associated with increased risk of all-cause mortality, as well as mortality due to malignant neoplasms, heart disease, cerebrovascular diseases, respiratory diseases, diabetes mellitus, and others in the general population. Additionally, since RAR was assessed from routine laboratory test, it could be recognized as a promising indicator that simply, reliably, and inexpensively accessible for identifying high-risk of mortality in clinical practice. Future studies are needed to validate these findings and investigate the causal relationship between social frailty and MCR risk.

## Data Availability

Data from NHANES are available at www.cdc.gov/nchs/nhis/index.htm, and data from UK Biobank are available on application at www.ukbiobank.ac.uk/register-apply.

## Acknowledgments

The data used in this research were obtained from the UK Biobank and NHANES. We would like to thank the workers, researchers, and participants involved in the UK Biobank and NHANES. This research has been conducted using the UK Biobank Resource under Application Number 103791.

## Author contributions

Zhang Hui and Hao Meng designed and conducted the research. Hao Meng, Li Xiangnan, Wang Shuming, Jiang Shuai, and Li Yi analyzed the data and performed statistical analyses. Zhang Hui, and Hao Meng drafted the manuscript. Zhang Hui and Hu Zixin supervised the study. Zhang Hui, Hao Meng, and Hu Zixin are primarily responsible for the final manuscript. All the authors read and approved the final manuscript.

